# Susceptibility and characteristics of infections in patients with glucocorticoid excess or insufficiency: The ICARO tool

**DOI:** 10.1101/2022.05.12.22275009

**Authors:** Marianna Minnetti, Valeria Hasenmajer, Emilia Sbardella, Francesco Angelini, Chiara Simeoli, Nicola Di Paola, Alessia Cozzolino, Claudia Pivonello, Dario de Alcubierre, Sabrina Chiloiro, Roberto Baldelli, Laura De Marinis, Rosario Pivonello, Riccardo Pofi, Andrea M. Isidori

## Abstract

**Objective:** Registry data show that Cushing’s syndrome (CS) and adrenal insufficiency (AI) increase mortality rates associated with infectious diseases. Little information is available on susceptibility to infections, especially milder forms that do not require hospitalization or prescription drugs. This study aimed to investigate infectious diseases in patients with glucocorticoid disorders through the development of a specific tool.

**Methods:** We developed and administered the ICARO questionnaire addressing infectious events over a 12-month observation period to 1017 outpatients referred to four University Hospitals between 2018 and 2019. The ICARO questionnaire showed good test-retest reliability. The odds of infection were estimated after adjustment for potential confounders and collated into the ICARO score, reflecting the frequency and duration of infections.

**Results:** 75 patients were diagnosed with hypercortisolism and 135 with hypocortisolism; 237 were excluded for uncontrolled disease. The remaining 570 served as controls. Compared to controls, hypercortisolism was associated with higher odds of respiratory (2.3) and urinary tract infections (4.1), mycoses (3.3) and flu (2.1). Subgroup analysis confirmed an increased risk of UTI (3.7) and flu (3.2) in mild hypercortisolism. Post-dexamethasone cortisol levels correlated with the ICARO score. Hypocortisolism also exhibited higher odds of mycoses (2.3), gastrointestinal (2.2) and urinary tract infections (2.5), independently of any glucocorticoid replacement dose.

**Conclusions:** ICARO is the first of its kind questionnaire developed to address susceptibility to infections in patients with glucocorticoid disorders and revealed the high prevalence and features of self-reported infections. ICARO could be a valuable tool for monitoring infections in various clinical settings.

## INTRODUCTION

The advent of the novel coronavirus disease (COVID-19) in early 2020 changed the world. It led the scientific community, including endocrinologists [1, 2], to refocus on the importance of infectious diseases in endocrine patients. COVID-19 has shown that mild infections may be asymptomatic in some subjects but can turn into a life-threatening disease in more susceptible patients, in whom systemic complications determine the outcome. Endocrinologists thus know that endocrine patients at risk of complications from infectious diseases need special care [3-7], but currently have no dedicated tools that can objectively estimate this risk.

Glucocorticoids (GCs) have a profound effect on host response, both stimulating and inhibiting the immune system and altering susceptibility to infections [8]. Overt GC excess, known as Cushing’s Syndrome (CS), has been associated with increased comorbidities, including cardiovascular disorders, diabetes and immune suppression [9-11], while infectious diseases are considered one of the main causes of death in patients with active CS [12-15]. To date, only a few studies have reported the prevalence of infections during the active phase of CS, estimated as between 21–51% [9]. However, these data were skewed by the inclusion of ectopic CS, a more severe condition with oncological confounders [16-19] and, importantly, only refer to infections leading to hospitalization. It is therefore impossible to discern whether CS increases susceptibility to infections or increases their severity, leading to increased hospitalization. The little data available on pituitary Cushing’s Disease (CD) mainly reported mortality. While possible autonomous cortisol secretion (pACS) from adrenal incidentalomas [20] and an increased incidence of cardiovascular and metabolic complications are reported in the setting of mild hypercortisolism, there is a lack of data on infectious diseases [21-23]. Finally, there are no data on the commonest, mild infections, which endocrinologists often face in outpatient clinics.

On the other side of the coin, adrenal insufficiency (AI), or hypocortisolism, has also been associated with premature mortality, in some cases ascribed to adrenal crisis triggered by infectious diseases [24-29]. Registry data reveal that AI patients seem to use more antimicrobial agents and are more often hospitalized for infections than matched controls [30, 31]. Again, less severe infections that do not require hospitalization or prescription drugs might be missed by public health or insurance registries – yet these events are still capable of triggering complications and affecting quality of life [32-34]. A recent trial revealed a higher cumulative incidence of different infections in AI patients in comparison with healthy controls [35].

To date, no large cohort studies have investigated the prevalence, type and severity of infections in patients with hypercortisolism (endogenous CS and pACS) or primary (PAI) and secondary (SAI) adrenal insufficiency. The aim of this study is to customize a new tool (a self-completed questionnaire) to assess susceptibility to infections in endocrine patients with various degrees of GC excess or deficiency in real-life outpatient settings.

## SUBJECTS AND METHODS

Between January 2018 and June 2019, we developed a questionnaire addressing infectious events in the previous 12 months and administered it to 1017 consecutive participants aged between 18 and 80 years attending the Endocrinology outpatient clinics at Sapienza University of Rome, Federico II University of Naples, Catholic University of Rome or San Camillo-Forlanini Hospital of Rome. All patients were asked to complete a self-reported 30-item questionnaire on the type, frequency, duration and treatment of any infectious diseases they had had in the previous 12 months. All participants gave their informed verbal consent to completion of the questionnaire and underwent a full medical interview. The study was performed in accordance with the principles of the Declaration of Helsinki and patient enrolment was authorized by the Sapienza University of Rome Ethics Committee (reference numbers 4945 and 7279). Questionnaires were administered confidentially, and the results were recorded under an anonymous alphanumeric code for each patient.

### The questionnaire

The questionnaire used is a structured tool based on the German National Cohort (GNC) Questionnaire [36, 37] and the immune system assessment questionnaire (ISAQ) [38]. Both of these questionnaires have been validated as self-administered tools with a good test-retest reliability. Like them, our tool, named ICARO (InfeCtions in pAtients with endocRinOpathies), contains items on the type of infections occurring over the previous 12 months. These investigate upper respiratory tract infections (URTIs: colds and infections of the sinus, tonsils, middle ear, throat and larynx), lower respiratory tract infections (LRTIs: bronchitis and pneumonia), gastrointestinal infections (“stomach flu”) (GIIs), two clusters of skin and soft tissue infections (SSTIs-1: herpes labialis, genital herpes, warts, herpes zoster, and conjunctivitis, and SSTIs-2: boils, abscesses, and styes), sexually transmitted infections (STIs), and urinary tract infections (UTIs). The questionnaire also investigated the frequency (none, 1–2 times; 3–4 times; 5–6 times, more than 6 times/year), duration (less than one week, 1-2 weeks, 2-3 weeks, more than 3 weeks), and treatment (antibiotics, antivirals, antifungals) of these infections [36, 37]. The ICARO score was developed as quantitative measure to test whether the type or severity of the GC imbalance had any impact on the frequency and duration of infections. Frequency was weighted in accordance with the ISAQ validation [38]: a negative response was weighted as 0.5, while ordinal frequency answers were reduced to binary items (i.e., 1-2 times and 3 times or more), with a weighting of 2 and 4, respectively. The duration of infections was also taken into account: the answer “less than one week” was weighted as 0, while other answers were again reduced to binary items (i.e., from one to two weeks and more than two weeks), with a weighting of 2 and 4, respectively. The arithmetic sum of the weights attributed to each item determined the total score (ICARO score). The range of possible scores is 4.5 to 64, with a higher score indicating a higher frequency and duration of infections. The observation time (12 months), derived from the GNC and ISAQ questionnaires, was chosen because it avoids biases due to seasonal variations in infection rates [36].

Compared to the GNC, which is designed for the general population, we added two items addressing mycosis (MYC) and flu and omitted items not directly related to infections, as ICARO is designed for use in outpatient clinics, where the patient’s medical history (PMH) is available. In fact, ICARO is designed to quantify susceptibility to infection independently of the underlying endocrine disorder, making it a single tool that can be offered to different types of endocrine patients; the inclusion of specific PMH questions would have limited this opportunity. We also added three items addressing hospital admission, absence from work, and vaccination (Supplementary data 1).

Demographic and clinical data were collected from patients’ medical records. The questionnaire was translated into Italian, and its translation validated for comprehensibility. Although the questionnaire was self-reported, a clinician was always available for clarifications and to ensure all items were answered. For questionnaire validation, test-retest reliability was assessed in a small group of patients (n=20) that were asked to complete the questionnaire twice within 30 days (range 8-28 days). Unweighted Cohen’s kappa coefficient was calculated for each question with optimal concordance for all the questionnaire items (κ≥0.85). Lastly, to assess the test-retest reliability of the questionnaire score, Chronbach’s alpha was calculated (α=0.989) and interclass correlation (ICC) was evaluated with a two-way mixed models assuming absolute agreement. The ICC of single measurements was 0.978 (95% CI 0.937-0.992). Overall, the test-retest reliability of the questionnaire was excellent. As an additional validating step, we had previously tested recall bias in a shorter version of the ICARO questionnaire used in the DREAM trial [35], in which recollected data from the questionnaire were paired with prospectively collected adverse events. There was a strong correlation between URTIs observed as adverse events and URTIs self-reported in the questionnaire (r= 0.986; *p*<0.001), again supporting the questionnaire’s reliability.

### Subjects

The groups of interest were subjects with endogenous hypercortisolism or hypocortisolism. The hypercortisolism group consisted of patients with endogenous CS and pACS. CS patients were subjects with pituitary ACTH-dependent or ACTH-independent hypercortisolism and typical laboratory findings and clinical signs and symptoms of CS [39, 40]. The questionnaire was administered to CS patients with typical biochemical and clinical signs of excess cortisol for more than a year and before commencement of any treatment for excess cortisol. Patients with adrenal adenoma, cortisol levels between 51 and 138 nmol/L after 1 mg overnight dexamethasone test (1 mg-DST) and a non-CS phenotype were considered as pACS, as reported in the recent European Society of Endocrinology guidelines [20]. The hypocortisolism group comprised patients with PAI and SAI diagnosed according to guidelines and treated with GCs plus daily doses of fludrocortisone as needed [41]. All patients were clinically stable, with no medication changes for their GC disorder in the 12 months before enrolment. All patients with hypocortisolism received an emergency kit and training in how to avoid adrenal crisis.

The control group consisted of consecutive endocrine patients attending the outpatient clinics for non-functioning adrenal adenomas, non-functioning pituitary adenomas without pituitary insufficiency, microprolactinomas, thyroid nodules, treated hypothyroidism, or well-controlled diabetes mellitus. The exclusion criteria were: uncontrolled diabetes (Hba1c >7.5%), ectopic CS, clinical or laboratory signs of significant haematologic, cerebral, cardiovascular, respiratory, hepatobiliary, pancreatic or kidney diseases, HIV, autoimmune polyendocrinopathy-candidiasis-ectodermal dystrophy (APECED), current malignant cancers (including thyroid cancer), severe psychiatric conditions, immunosuppressant or corticosteroid treatment for rheumatological, pneumological, or allergic conditions, pregnancy, or hyperthyroidism/hypothyroidism not adequately controlled at screening. Age, sex, smoking status, overweight/obesity, diabetes mellitus diagnosis, reproductive function, past medical history, disease duration and medications were recorded for all subjects. In the hypocortisolism group, steroids were converted into daily hydrocortisone equivalent doses. Recruitment was set to a sample size of 1000, considered by Comfrey and Lee (1992) as ‘excellent’ for subsequent analyses [42].

### Statistical analysis

Population characteristics were summarized as counts and percentages for categorical variables and as means and standard deviations for continuous variables. Distribution of continuous variables was tested with the Shapiro–Wilk test; linearity was established by visual inspection of a scatterplot. Categorical variables were expressed as percentage and frequency; continuous variables were reported as mean and 95% confidence interval (95% CI) or median and interquartile range (IQR) (25th–75th percentile). The association between categorical variables was determined using the Chi-square test with continuity correction. For multiple ordinal variables a post hoc analysis involving pairwise comparisons using multiple z-tests of two proportions with a Bonferroni correction was performed. Unpaired Student’s T test and Mann–Whitney test were applied to compare groups, as appropriate. Multivariate logistic regression was used to estimate the odds of having an infection, with adjustment for potential confounders (age, sex, diabetes, menopause/hypogonadism, obesity). The diagnostic performance of the ICARO score was evaluated using areas under the receiving-operating characteristic curve (AUC of ROC). Spearman’s correlation was performed to test the correlation between infection score and both GCs dose and duration. Regression analysis was performed to estimate GC contributions to ICARO score variability. Because the questionnaire covered an observation time of 12 months, we considered dichotic comorbidities (with a duration of at least one year) as confounders, as opposed to variables that could change drastically over the year (such as body mass index, Hba1c or arterial blood pressure). The sensitivity analysis was performed first by excluding patients with obesity or diabetes from the entire cohort, then by analysing only the cohort of subjects with obesity or diabetes. In view of the multiple independent tests, p-values were adjusted using the false discovery rate (FDR) method (Benjamini and Hochberg) [43]. FDR-corrected p <0.05 was considered significant. Statistical analyses were conducted using the statistical software IBM SPSS statistics 20.

## RESULTS

Of the 1017 patients screened for study inclusion, 222 did not meet the inclusion criteria and 15 did not complete the entire questionnaire; 237 patients were therefore excluded from the analysis. The remaining 780 patients were enrolled (**Figure 1**). Of these, 210 patients had GC disorders and 570 patients were included in the control group. Among patients with GC disorders, 75 had a diagnosis of hypercortisolism (43 met the criteria for CS and 32 for pACS) and 135 of hypocortisolism (32 PAI and 103 SAI) (**Figure 1**).

**Figure 1.**
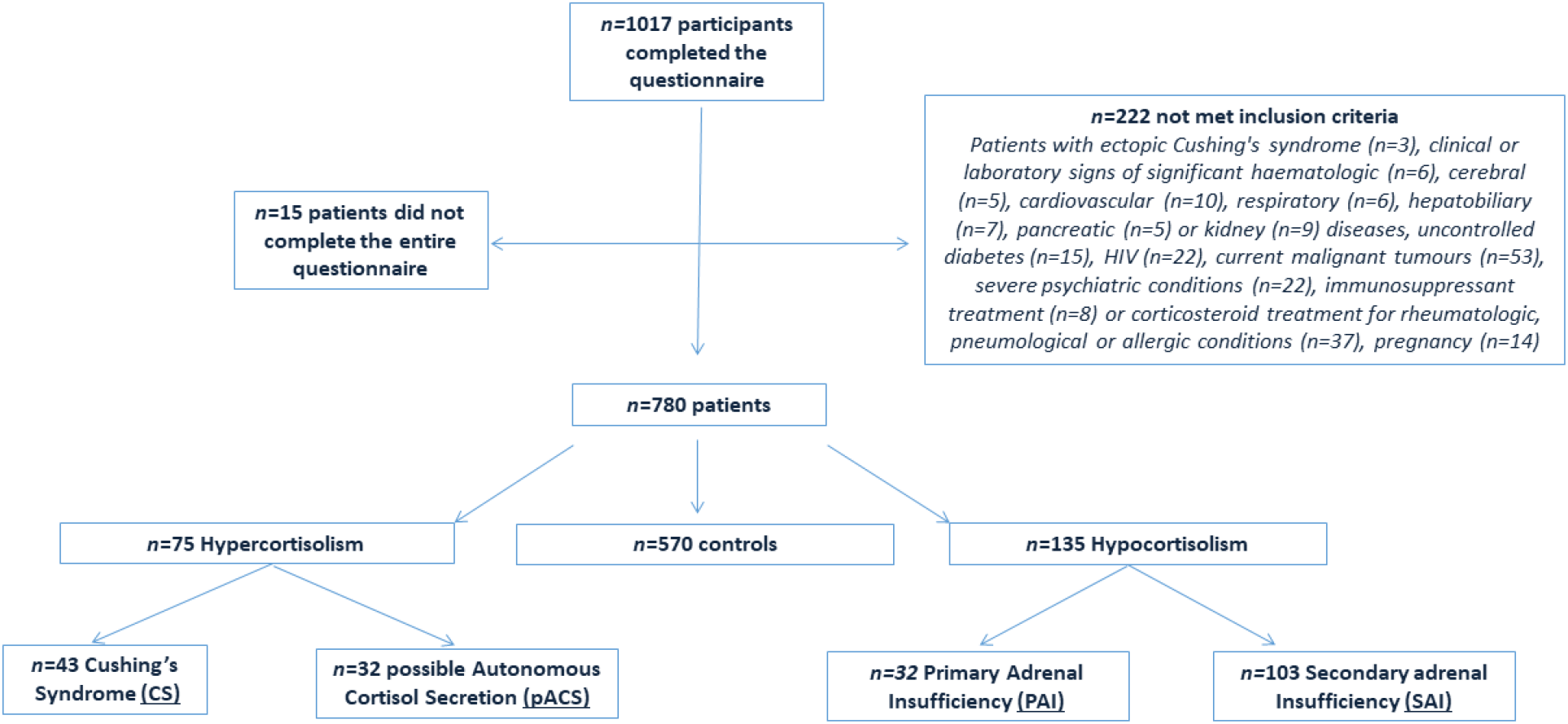
Study flow diagram.

Table 1 and Table 2 describe the clinical characteristics of the study population. The distribution of the total infection frequency score, which measures the cumulative weighted incidence of different infections over 12 months, is reported in **Figure 2**. Hospitalization was required for just 1% and prescription medicines for 48% of infections in patients with GC disorders.

**Table 1.** Clinical characteristics in the study population. pACS, possible autonomous cortisol secretion, PAI primary adrenal insufficiency. Categorical variables are expressed as frequency and percentages. Continuous variables are expressed as mean and standard deviation. \**p* values for comparisons with controls.

|  | All subjects<br>(n=780) | Controls<br>(n=570) | Hypercortisolism<br>(n=75) | *P* * | CS<br>(n=43) | *P* * | pACS<br>(n=32) | *P* * |
| --- | --- | --- | --- | --- | --- | --- | --- | --- |
| Age (years) | 49.9±15.8 | 49.1±16.3 | 54.2±14.2 | 0.005 | 46.2±12.1 | 0.243 | 65±8.3 | <0.001 |
| Females | 54% (424) | 52% (294) | 81% (61) | <0.001 | 86% (37) | <0.001 | 75% (24) | 0.065 |
| Diabetes | 9% (71) | 7% (38) | 23% (17) | <0.001 | 33% (14) | <0.001 | 9% (3) | 1.000 |
| Obesity | 16% (124) | 14% (78) | 33% (25) | <0.001 | 39.5% (17) | <0.001 | 25% (8) | 0.514 |
| Menopause/<br>Hypogonadism | 49% (382) | 47% (266) | 61% (46) | 0.033 | 46.5% (20) | 0.914 | 81% (26) | 0.002 |
| Smoking status | 19% (150) | 17% (99) | 24% (18) | 0.343 | 23% (10) | 0.443 | 25% (8) | 0.389 |

**Table 2.** Prevalence, recurrence, duration, therapy of infectious diseases and infection frequency score in patients with hypercortisolism and in the subgroups, possible autonomous cortisol secretion (pACS) and primary adrenal insufficiency (PAI). Categorical variables are expressed as percentages and frequencies. Continuous variable is expressed as median (25th–75th percentile). \**p* values for comparisons with controls.

|  | All subjects<br>(n=780) | Controls<br>(n=570) | Hypocortisolism<br>(n=135) | *P* * | PAI<br>(n=32) | *P* * | SAI<br>(n=103) | *P* * |
| --- | --- | --- | --- | --- | --- | --- | --- | --- |
| Age (years) | 49.9±15.8 | 49.1±16.3 | 50.5±14.6 | 0.460 | 50.3±15.1 | 0.699 | 50.6±14.5 | 0.421 |
| Females | 54% (424) | 52% (294) | 51% (69) | 0.998 | 53% (17) | 1.000 | 16.5% (17) | 0.829 |
| Diabetes | 9% (71) | 7% (38) | 12% (16) | 0.190 | 12.5% (4) | 0.366 | 12% (12) | 0.099 |
| Obesity | 16% (124) | 14% (78) | 16% (21) | 0.850 | 12.5% (4) | 1.000 | 16.5% (17) | 0.436 |
| Menopause/<br>Hypogonadism | 49% (382) | 47% (266) | 52% (70) | 0.968 | 47% (15) | 1.000 | 53% (55) | 0.207 |
| Smoking<br>status | 19% (150) | 17% (99) | 24% (33) | 0.263 | 19% (6) | 1.000 | 27% (27) | 0.188 |

**Figure 2.**
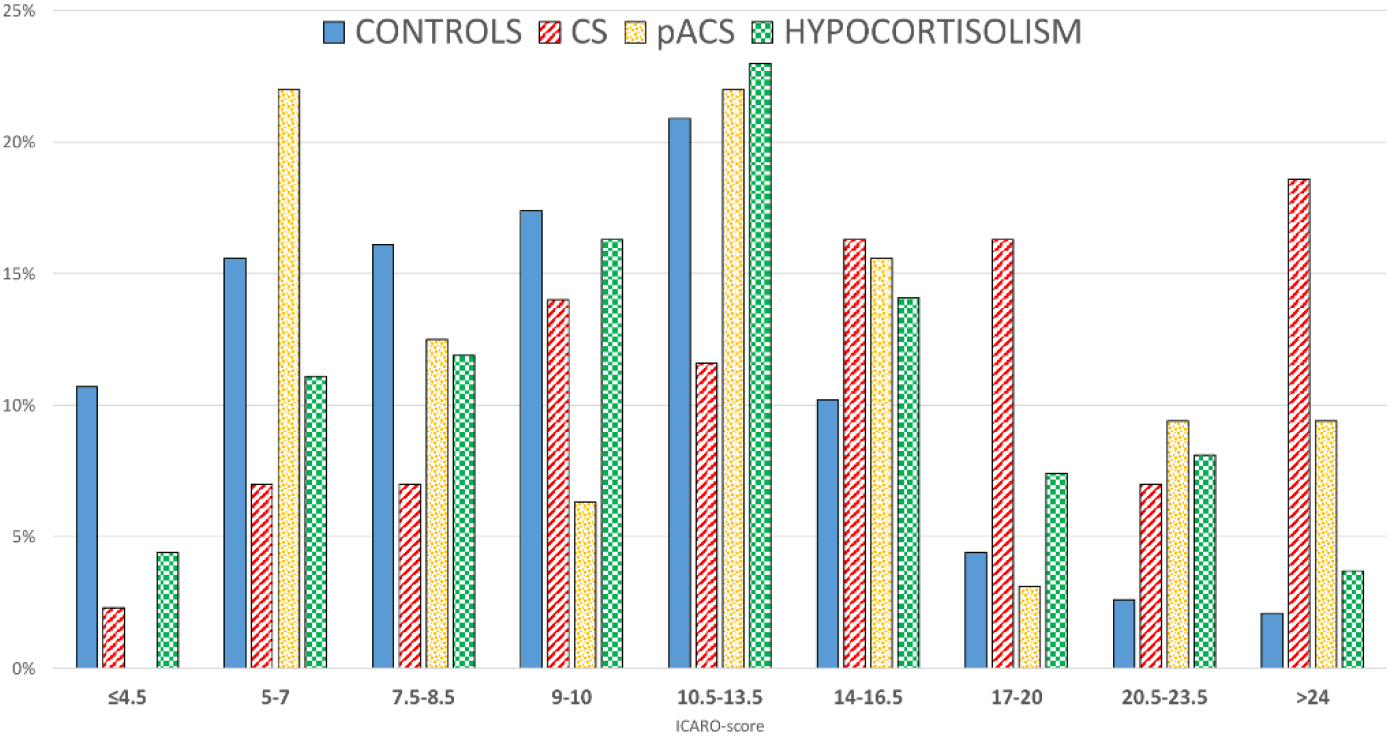
Distribution of ICARO score in controls (blue), patients with Cushing’s syndrome (CS) (red lines), possible autonomous cortisol secretion (pACS) (orange dots) and hypocortisolism (green squares).

### Hypercortisolism: all subjects

This group included 75 patients: 43 with overt CS (38 pituitary, 5 adrenal) and 32 with pACS. Median 1 mg-DST was 92 nmol/l (IQR 68.9-137). Multivariate logistic regression analysis, corrected for age, diabetes, obesity, menopause/hypogonadism and sex, showed that patients with hypercortisolism had higher odds of developing more LRTIs [OR 2.3 (95% CI, 1.2 to 4.3); *p*=0.049)], MYC [OR 3.3 (95% CI, 1.8 to 6.1); *p*=0.033], UTIs [OR 4.1 (95% CI, 2.4 to 7.1); *p*=0.011] and flu [2.9 (95% CI, 1.7 to 5); *p*=0.011)] in the year preceding questionnaire administration (**Figure 3**).

**Figure 3.**
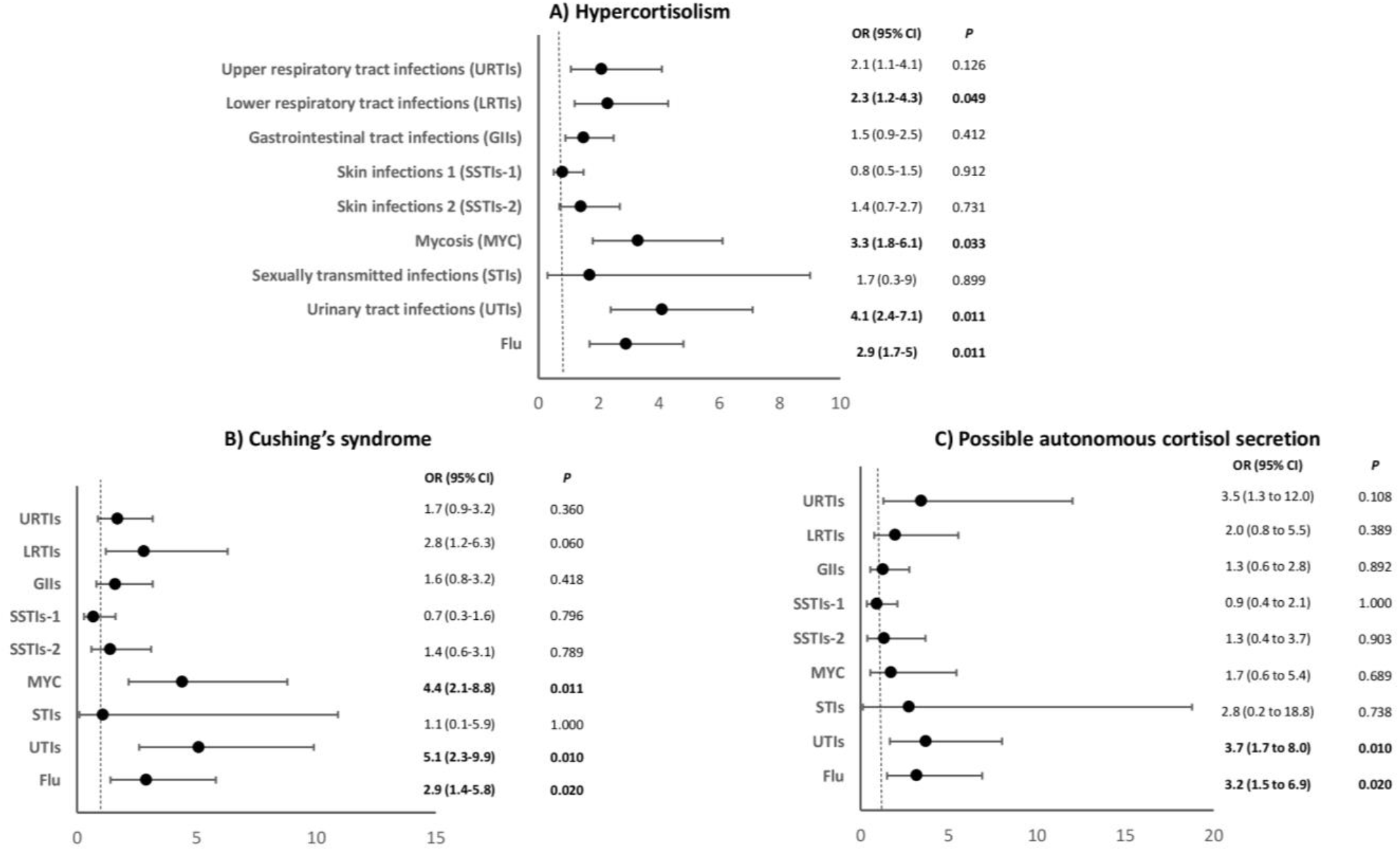
Odds of having an infectious disease in the 12 months prior to questionnaire administration, adjusted for age, sex, diabetes, obesity, and menopause/hypogonadism in patients with hypercortisolism (A), Cushing’s syndrome (B) and possible autonomous cortisol secretion (C) compared with controls.

The proportion of subjects reporting at least one infectious episode was higher in this cohort than in controls for LRTIs (24% vs 11%; *p*=0.016), MYC (31% vs 11%; *p*=0.025), UTIs (49% vs 15%; *p*=0.023) and flu (64% vs 42%; p=0.026) (Table 3). Patients with hypercortisolism also reported a longer duration of infections, specifically for GIIs (*p*=0.029), and MYC (*p*=0.025) (Supplementary Table 1). Moreover, patients with hypercortisolism also had a higher ICARO score than controls (11.5 vs 9.5; *p*=0.021).

**Table 3.** Prevalence and therapy of infectious diseases and infection frequency score in patients with hypercortisolism. Categorical variables are expressed as percentages and frequencies. Continuous variable is expressed as median (25th–75th percentile). *p values for comparisons with controls. §p values for comparisons between CS and pACS. After correction for multiple testing, *p* <0.05 was considered a statistically significant difference (bold).

| | Controls<br>(n=570) | Hypercortisolism<br>(n=75) | $p^*$ | CS<br>(N=43) | $p^*$ | pACS<br>(n=32) | $p^*$ | $p^§$ |
| --- | --- | --- | --- | --- | --- | --- | --- | --- |
| Upper respiratory tract infections (URTis) | 75% (429/570) | 84% (63/75) | 0.367 | 81% (35/43) | 0.738 | 87.5% (28/32) | 0.150 | 0.693 |
| Antimicrobials | 36% (154/429) | 44% (28/63) | 1.000 | 40% (14/35) | 0.742 | 46% (13/28) | 0.925 | 0.819 |
| Lower respiratory tract infections (LRTis) | 11% (64/570) | 24% (18/75) | **0.016** | 23% (10/43) | 0.086 | 25% (8/32) | 0.150 | 0.861 |
| Antimicrobials | 73% (47/64) | 83% (15/18) | 0.900 | 80% (8/10) | 0.892 | 87.5% (7/8) | 0.985 | 0.769 |
| Gastrointestinal tract infections (Giis) | 32.5% (185/570) | 41% (31/75) | 0.437 | 49% (21/43) | 0.112 | 31% (10/32) | 1.000 | 0.126 |
| Antimicrobials | 18% (33/185) | 13% (4/31) | 0.934 | 9.5% (2/21) | 0.732 | 20% (2/10) | 1.000 | 0.810 |
| Skin infections 1 (SSTis-1) | 25% (142/570) | 24% (18/75) | 1.000 | 23% (10/43) | 1.000 | 25% (8/32) | 1.000 | 0.861 |
| Antimicrobials | 39% (55/142) | 72% (13/18) | 0.064 | 90% (9/10) | **0.019** | 50% (4/8) | 1.000 | 0.065 |
| Skin infections 2 (SSTis-2) | 15% (88/570) | 19% (14/75) | 0.895 | 23% (10/43) | 0.459 | 12.5% (4/32) | 1.000 | 0.377 |
| Antimicrobials | 27% (24/88) | 21% (3/14) | 1.000 | 20% (2/10) | 0.034 | 25% (1/4) | 1.000 | 0.837 |
| Mycosis (MYC) | 11% (63/570) | 31% (23/75) | **0.025** | 42% (18/43) | **0.018** | 16% (5/32) | 0.899 | **0.029** |
| Antimicrobials | 81% (51/63) | 74% (17/23) | 0.935 | 83% (15/18) | 1.000 | 40% (2/5) | 0.360 | 0.169 |
| Sexually transmitted infections (STIs) | 2% (9/570) | 3% (2/75) | 1.000 | 2% (1/43) | 0.951 | 3% (1/32) | 1.000 | 1.000 |
| Antimicrobials | 56% (5/9) | 50% (1/2) | 1.000 | 100% (1/1) | 0.752 | 0% (0/1) | 1.000 | 1.000 |
| Urinary tract infections (UTIs) | 15% (86/570) | 49% (37/75) | **0.023** | 53.5% (23/43) | **0.015** | 44% (14/32) | **0.013** | 0.548 |
| Antimicrobials | 76% (65/86) | 59.5% (22/37) | 0.358 | 74% (17/23) | 1.000 | 36% (5/14) | 0.062 | 0.051 |
| Flu | 42% (240/570) | 64% (48/75) | **0.026** | 67% (29/43) | **0.021** | 59% (19/32) | 0.082 | 0.634 |
| Hospitalization due to infectious disease | 1% (5/570) | 3% (2/75) | 0.812 | 2% (1/43) | 0.736 | 3% (1/32) | 0.977 | 0.832 |
| Vaccination | 20% (112/570) | 27% (20/75) | 0.571 | 21% (9/43) | 1.000 | 34% (11/32) | 0.260 | 0.299 |
| Absence from work due to infectious disease | 10% (56/570) | 8% (6/75) | 0.927 | 12% (5/43) | 0.948 | 3% (1/32) | 0.757 | 0.362 |
| Infections frequency score | 9.5 (6-13) | 11.5 (8-15) | **0.021** | 16 (9-20.5) | **0.014** | 11.25 (7.5-16.5) | **0.034** | **0.049** |

A linear regression analysis showed that 1 mg-DST explained up to 22% of the ICARO score (*p*=0.001). ICARO score was in fact positively related to post-DST cortisol (r = 0.415, *p*< 0.001), but subgroup analysis in pACS and CS revealed that this was mainly driven by patients with CS (r = 0.581, *p* <0.001) rather than pACS (p=0.958) (**Figure 4**). In fact, the ICARO score was higher in CS (median 16, IQR 9-20) than pACS patients (median 11, IQR 7-16; *p*=0.049)

Sensitivity analysis revealed that the median frequency of infection (score) was significantly higher in patients with hypercortisolism compared to controls, even after excluding patients with diabetes or obesity from the entire cohort (11 vs 9.5; *p*=0.012). Similarly, a higher frequency of infection was also seen in this group when restricting the analysis to the subjects with diabetes or obesity (17.25 vs 9; *p*=0.011). No differences in prevalence, duration and treatment of infections were found between CS and pACS patients, except for prevalence of MYC, which was higher in CS patients (42% vs 16% respectively; *p*=0.029).

**Figure 4.**
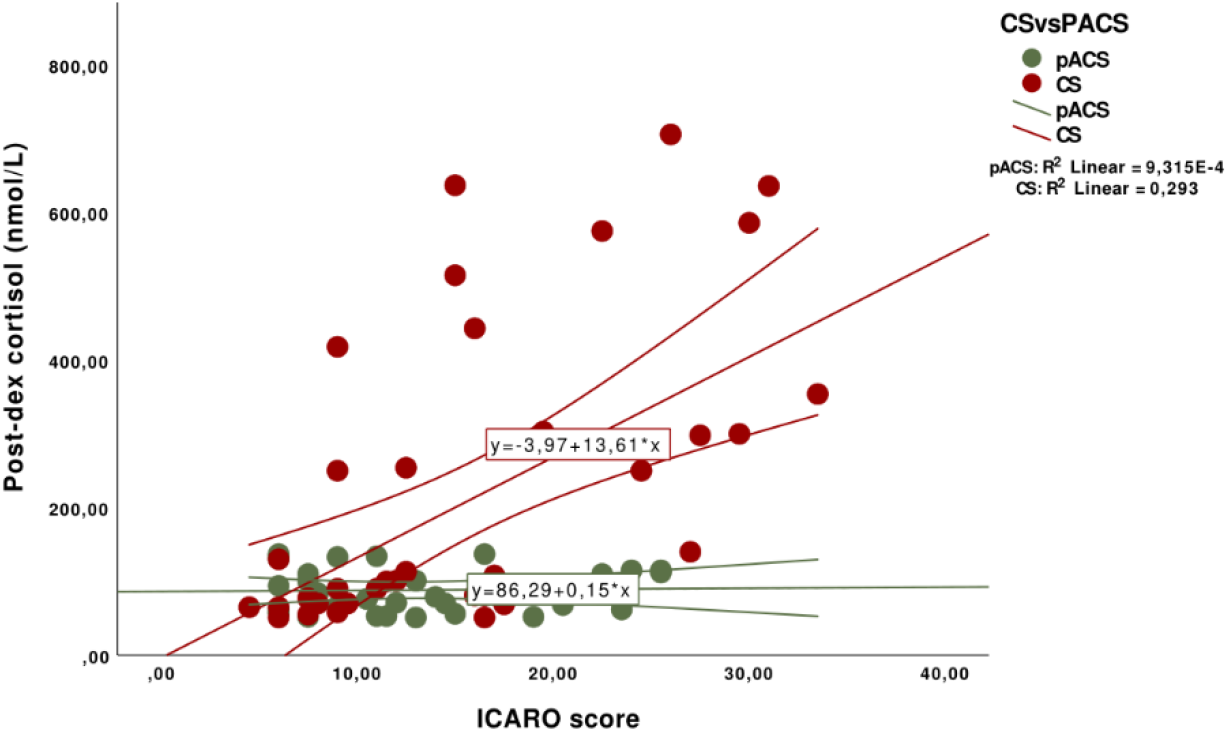
Correlation between post-DST cortisol levels and ICARO score in patients with Cushing’s syndrome (CS) and possible autonomous cortisol secretion (pACS).

### Cushing’s Syndrome

The 43 CS patients had a median 1mg-DST of 101 nmol/l (IQR 77-300) and typical clinical signs of CS. The multivariate logistic regression showed that these patients had higher adjusted odds of developing MYC [OR 4.4 (95% CI, 2.1 to 8.8); *p*=0.011], UTIs [OR 5.1 (95% CI, 2.3 to 9.9); *p*=0.010] and flu [OR 2.9 (95% CI, 1.4 to 5.8); *p*=0.020] compared to controls (**Figure 3**). Patients with CS were also more likely to report at least one episode of MYC (42% vs 11%; *p*=0.018) and UTIs (53.5% vs 15%; *p*=0.015) in the timeframe covered by the questionnaire (Table 3).

Patients with CS also reported a longer duration of the infectious episode, specifically for GIIs (p=0.037), SSTIs-2 (p=0.034) and MYC (*p*=0.016) (Supplementary Table 1), than the controls. They also took antimicrobials to treat SSTIs-1 significantly more often than controls (90% vs 39% respectively; p=0.019) (Table 3). Finally, they had a higher infection score than controls (16 vs 9.5 respectively; p=0.014). ROC analyses [AUC 0.736 (0,699 to 0,770); p<0.001] showed that an ICARO score >14.5 discriminated CS from controls (sensitivity 58%, specificity 85%, negative predictive value 96%) (**Figure 5**).

**Figure 5.**
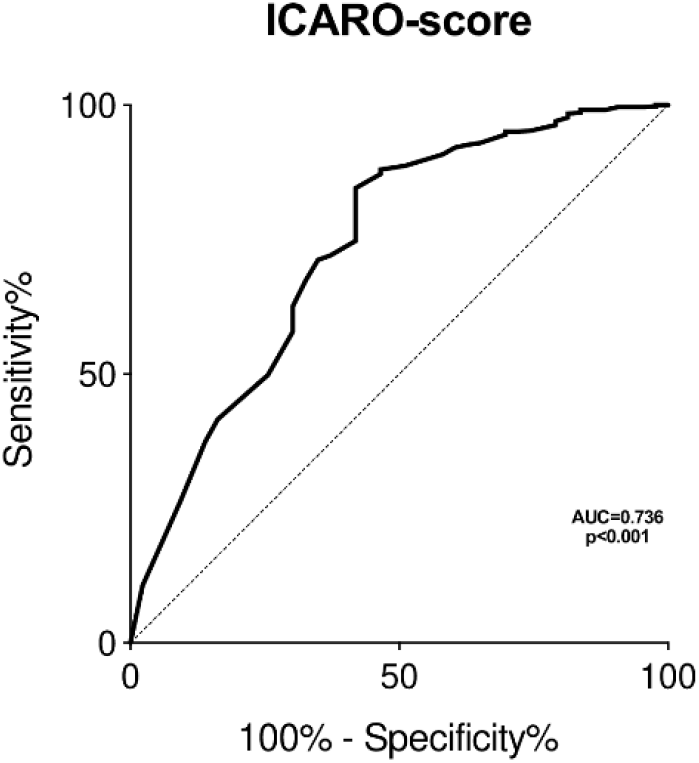
ROC curve analysis of the ICARO score ability to predict Cushing’s syndrome.

### Possible autonomous cortisol secretion

The 32 patients with pACS showed a median 1mg-DST of 81.5 nmol/l (59-114). In this group, the multivariate logistic regression showed an increased adjusted odds of developing recurrent UTIs [OR 3.7 (95% CI, 1.7 to 8.0); *p*=0.010)] and flu [OR 3.2 (95% CI, 1.5 to 6.9); *p*=0.020] compared to controls (**Figure 3)**. These patients were also more likely to report at least one episode of UTI (44% vs 15%; *p*=0.013) (Table 3). They also had a higher ICARO score than the controls (11.25 vs 9.5; *p*=0.012).

### Hypocortisolism: all subjects

AI patients presented higher adjusted odds of developing frequent GIIs [OR 2.2 (95% CI, 1.5 to 3.3); *p*=0.009], MYC [OR 2.3 (95% CI 1.4 to 3.8; *p*=0.009)], and UTIs [OR 2.5 (95% CI, 1.6 to 3.9); *p*<0.008] compared with controls (**Figure 6**).

**Figure 6.**
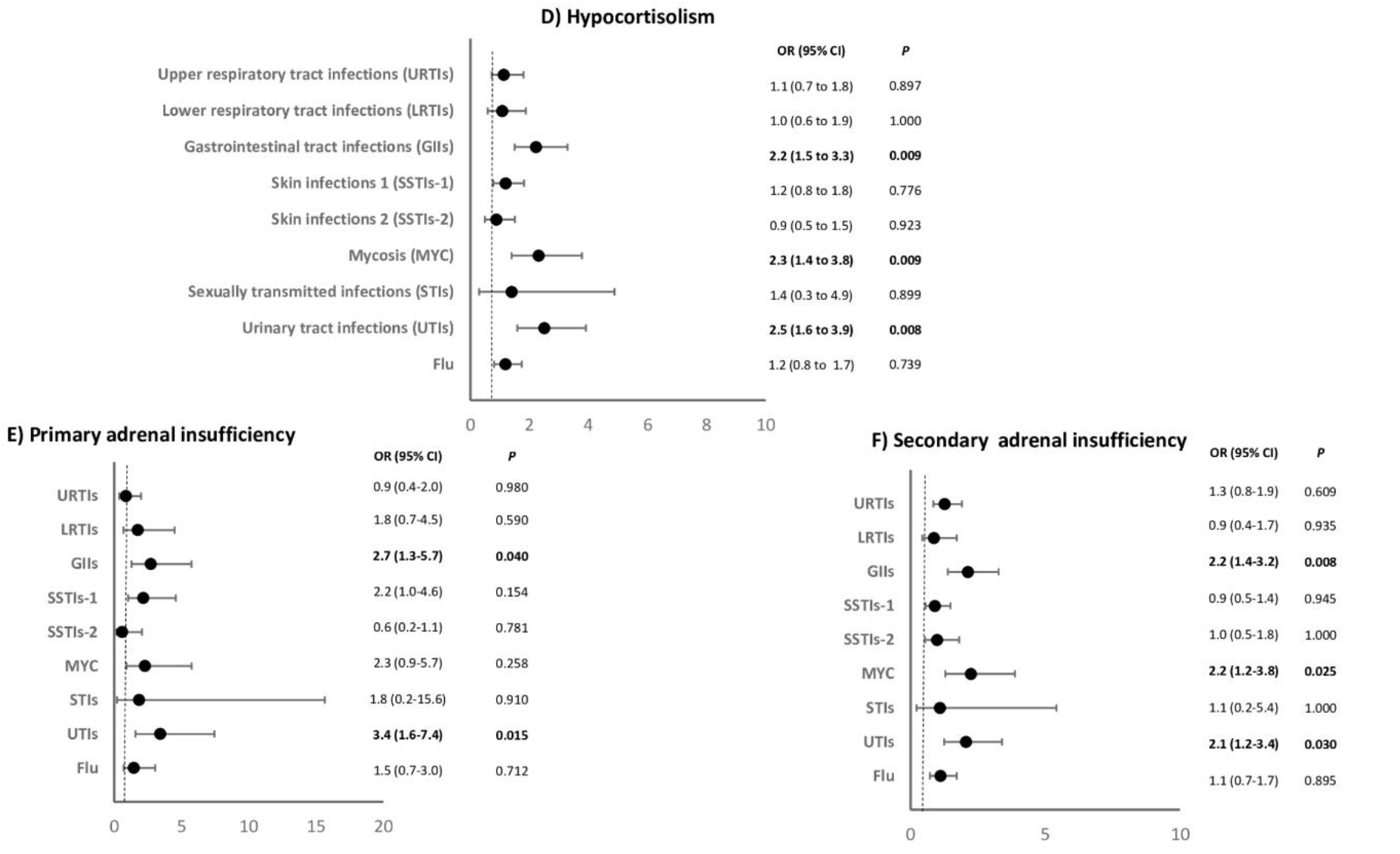
Odds of having an infectious disease in the 12 months prior to questionnaire administration, adjusted for age, sex, diabetes, obesity, and menopause/hypogonadism in patients with adrenal insufficiency (D), primary adrenal insufficiency (E) and secondary adrenal insufficiency (F) compared with controls.

Patients with AI were more likely to experience at least one episode of GIIs (50% vs 32.5%; *p*=0.013), MYC (22% vs 11%, *p*=0.012), UTIs (30% vs 15%; *p*=0.030) and flu (64% vs 42%; *p*=0.030) as well as long-lasting LRTIs (*p*=0.033) (supplementary Table 2) than the controls. All AI patients were diagnosed at least one year before enrolment and had a stable replacement therapy with a median hydrocortisone equivalent dose of 20 mg/day (IQR 20-30). There was no significant difference in the replacement dose between subgroups (20 mg/day, IQR 20-25, in PAI patients and 25 mg/day, IQR 15-30, in SAI patients, p=0.968) or in the median GC therapy duration (4.5 years, IQR 2-10, for PAI patients and 4 years, 2-9, for SAI patients, *p*=0.707). A binomial logistic regression adjusted for these confounding factors excluded any major role of GC dose or duration of therapy as risk factors for GIIs, MYC or UTIs. Patients with AI had a significantly higher ICARO score than the controls (11.5 vs 9.5; *p*=0.038), even after controlling for daily GC dose and duration and excluding subjects with obesity and diabetes. When the analysis was restricted to the latter subgroup, the frequency of infections was again higher than in the controls (10.5 vs 9; *p*<0.039). No differences in prevalence, duration or ICARO score were found between primary and secondary AI.

### Primary adrenal insufficiency

The PAI subgroup showed higher adjusted odds of developing recurrent GIIs [OR 2.7 (95% CI 1.3 to 5.7); *p*=0.040] and UTIs [OR 3.4 (95% CI 1.6 to 7.4); *p*=0.015], compared to controls (**Figure 6**) and were more likely to report at least one episode of UTI (38% vs 15%; *p*=0.015) and flu (67% vs 42%; *p*=0.031) (Table 4). The average URTI duration was higher in PAI patients compared to controls (*p*=0.036) (Supplementary Table 2).

**Table 4.**
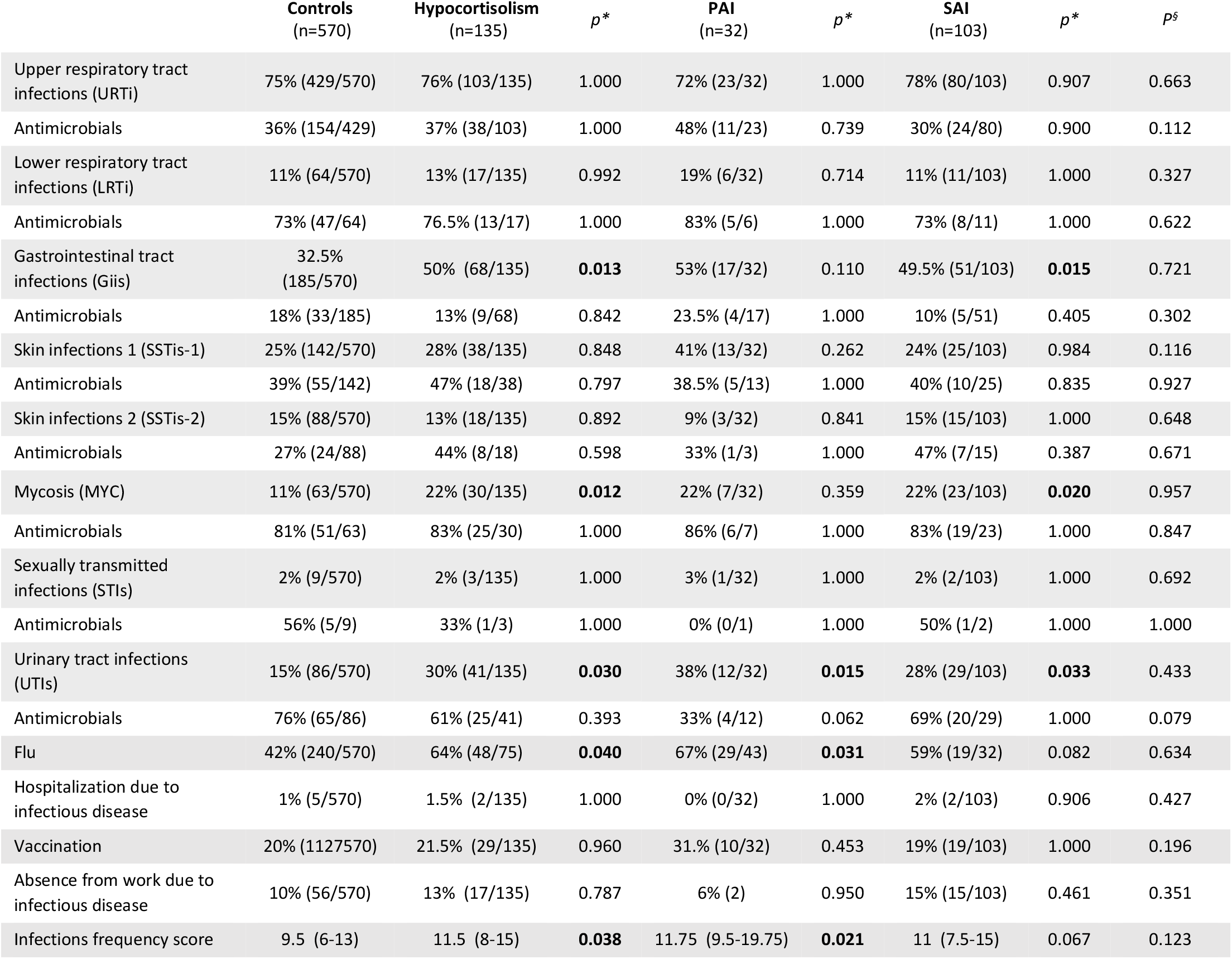
Prevalence, therapy of infectious diseases and infection frequency score in patients with hypocortisolism. Categorical variables are expressed as percentages and frequencies. Continuous variable is expressed as median (25th–75th percentile). \**p* values for comparisons with controls. ^§^*p* values for comparisons between PAI and SAI. After correction for multiple testing, *p* <0.05 was considered a statistically significant difference (bold).

|  | Controls<br>(n=570) | Hypocortisolism<br>(n=135) | *p* * | PAI<br>(n=32) | *p* * | SAI<br>(n=103) | *p* * | *p*<sup>§</sup> |
| --- | --- | --- | --- | --- | --- | --- | --- | --- |
| Upper respiratory tract infections (URTi) | 75% (429/570) | 76% (103/135) | 1.000 | 72% (23/32) | 1.000 | 78% (80/103) | 0.907 | 0.663 |
| Antimicrobials | 36% (154/429) | 37% (38/103) | 1.000 | 48% (11/23) | 0.739 | 30% (24/80) | 0.900 | 0.112 |
| Lower respiratory tract infections (LRTi) | 11% (64/570) | 13% (17/135) | 0.992 | 19% (6/32) | 0.714 | 11% (11/103) | 1.000 | 0.327 |
| Antimicrobials | 73% (47/64) | 76.5% (13/17) | 1.000 | 83% (5/6) | 1.000 | 73% (8/11) | 1.000 | 0.622 |
| Gastrointestinal tract infections (Giis) | 32.5% (185/570) | 50% (68/135) | **0.013** | 53% (17/32) | 0.110 | 49.5% (51/103) | **0.015** | 0.721 |
| Antimicrobials | 18% (33/185) | 13% (9/68) | 0.842 | 23.5% (4/17) | 1.000 | 10% (5/51) | 0.405 | 0.302 |
| Skin infections 1 (SSTis-1) | 25% (142/570) | 28% (38/135) | 0.848 | 41% (13/32) | 0.262 | 24% (25/103) | 0.984 | 0.116 |
| Antimicrobials | 39% (55/142) | 47% (18/38) | 0.797 | 38.5% (5/13) | 1.000 | 40% (10/25) | 0.835 | 0.927 |
| Skin infections 2 (SSTis-2) | 15% (88/570) | 13% (18/135) | 0.892 | 9% (3/32) | 0.841 | 15% (15/103) | 1.000 | 0.648 |
| Antimicrobials | 27% (24/88) | 44% (8/18) | 0.598 | 33% (1/3) | 1.000 | 47% (7/15) | 0.387 | 0.671 |
| Mycosis (MYC) | 11% (63/570) | 22% (30/135) | **0.012** | 22% (7/32) | 0.359 | 22% (23/103) | **0.020** | 0.957 |
| Antimicrobials | 81% (51/63) | 83% (25/30) | 1.000 | 86% (6/7) | 1.000 | 83% (19/23) | 1.000 | 0.847 |
| Sexually transmitted infections (STIs) | 2% (9/570) | 2% (3/135) | 1.000 | 3% (1/32) | 1.000 | 2% (2/103) | 1.000 | 0.692 |
| Antimicrobials | 56% (5/9) | 33% (1/3) | 1.000 | 0% (0/1) | 1.000 | 50% (1/2) | 1.000 | 1.000 |
| Urinary tract infections (UTIs) | 15% (86/570) | 30% (41/135) | **0.030** | 38% (12/32) | **0.015** | 28% (29/103) | **0.033** | 0.433 |
| Antimicrobials | 76% (65/86) | 61% (25/41) | 0.393 | 33% (4/12) | 0.062 | 69% (20/29) | 1.000 | 0.079 |
| Flu | 42% (240/570) | 64% (48/75) | **0.040** | 67% (29/43) | **0.031** | 59% (19/32) | 0.082 | 0.634 |
| Hospitalization due to infectious disease | 1% (5/570) | 1.5% (2/135) | 1.000 | 0% (0/32) | 1.000 | 2% (2/103) | 0.906 | 0.427 |
| Vaccination | 20% (112/570) | 21.5% (29/135) | 0.960 | 31% (10/32) | 0.453 | 19% (19/103) | 1.000 | 0.196 |
| Absence from work due to infectious disease | 10% (56/570) | 13% (17/135) | 0.787 | 6% (2) | 0.950 | 15% (15/103) | 0.461 | 0.351 |
| Infections frequency score | 9.5 (6-13) | 11.5 (8-15) | **0.038** | 11.75 (9.5-19.75) | **0.021** | 11 (7.5-15) | 0.067 | 0.123 |

### Secondary adrenal insufficiency

These patients had increased adjusted odds of developing GIIs [OR 2.2 (95% CI, 1.4 to 3.2); *p*=0.008], MYC [OR 2.2 (95% CI, 1.2 to 3.8); *p*=0.025] and UTIs [OR 2.1 (95% CI, 1.2 to 3.4); *p*=0.030)] compared to controls (**Figure 6**). Moreover, they were also more likely to experience at least one episode of GIIs (49.5% vs 32.5%; p=0.015), MYC (22% vs 11%; *p*=0.020) and UTIs (28% vs 15%; *p*=0.033) (Table 4).

## DISCUSSION

This study reveals that the disruption of endogenous glucocorticoid levels, irrespective of the underlying cause, is associated with increased susceptibility to infections, documented by higher frequencies and longer durations of self-reported episodes measured with a new tool, ICARO, specifically developed to assess infection risk in endocrine patients. The immune impairment measured with the ICARO score correlates with the severity of hypercortisolism. Our evidence suggests that the increased mortality of patients with AI and CS is a consequence of increased susceptibility to mild infectious diseases that do not necessarily require hospitalization, but whose resolution is less efficient.

Although more than 50 years have passed since bacterial infections were first recognized as the leading cause of death in patients with CS [44], very few studies have addressed the epidemiology of infections in these patients [16-19]. Retrospective studies report infections mostly due to opportunistic pathogens, but are biased by the inclusion of ectopic CS [16-19]. Others reported infections as frequent comorbidities in up to 21% of CD patients, but without describing the type and severity [45]. A recent retrospective study on CD confirmed a 5-fold increased mortality due to infections [14]. The mechanisms through which GC excess affects the innate and adaptive immune system were recently reviewed [8, 9], but no studies have investigated the risk of common, mild, and/or recurrent infections in CS, probably because they evade surveillance strategies (hospital record/insurance registries) and there were no tools able of measuring their prevalence. Our study fills this gap and adds important information about the prevalence, odds and duration of the commonest infections in a cohort that included 75 patients with hypercortisolism. In these patients we measured a 2-to 4-fold increase in the odds for LRTIs, MYC, UTIs and FLU, compared to controls. This held true even after adjusting for age, sex, hypogonadism/menopausal state, obesity and diabetes, suggesting that the infection risk is independent of the commonest confounding factors. Moreover, we also revealed that GIIs and MYC can last longer in these patients. Awareness of the type, characteristics and duration of infections could have clinical implications, considering the atypical presentations of infectious disease in patients with CS, in whom hypercortisolism can mitigate clinical symptoms, leading to possible referral delays [8]. Interestingly, 1-mg-DST values are positively correlated with the ICARO score, arguing for a close relationship between the degree of hypercortisolism and the severity of infections. However, this was confirmed only for patients with overt CS. The ICARO questionnaire could therefore be useful during the diagnostic work-up of CS, and perhaps in pseudo-Cushing’s syndrome [46]; in our cohort a cut-off of <14.5 was specific enough to identify patients in whom CS is unlikely.

Our group of patients with hypercortisolism also included patients with adrenal incidentalomas and pACS [47]. Between 30% and 50% of patients with adrenal incidentalomas have biochemical evidence of low-grade autonomous cortisol release [20], associated with a worsened cardiovascular risk (hypertension, diabetes and obesity), a higher prevalence of cardiovascular events, greater bone loss, and a higher mortality rate [22, 47-49]. However, very little attention has been paid to the infection risk in patients with pACS. To the best of our knowledge, only one retrospective study has investigated patients with adrenal incidentaloma and pACS, reporting a higher infection-related mortality than in controls, but no data on prevalence and risk of infections were reported [50]. Our study is the first showing that even a low-grade cortisol excess results in a significant increase in some (UTIs and FLU), but not all, infections. Our subgroup analysis suggests different phenotypes in hypercortisolaemic patients: the increased odds for MYC in the whole cohort is mainly driven by the CS patients, while the increased frequency of UTIs and FLU is observed in both groups. Since the ICARO score seems to correlate with the severity of hypercortisolism, it could also offer an additional indication as whether to treat pACS patients with surgical or conservative management [20, 47].

Other than hypercortisolism, our results show that the prevalence and odds of contracting a greater number of GIIs, MYC and UTIs were also significantly higher in patients with hypocortisolism, when compared to controls, although the duration and treatment were similar.

Large retrospective studies have demonstrated that mortality is increased in chronic AI and is strongly corelated with infectious disease [24-26, 29, 51, 52]. According to Swedish registry data, one-third of patients with PAI and concomitant diabetes suffered from infections requiring hospital admission [25]. A recent British primary care retrospective database reported an increased risk of infections (LRTIs, UTIs and GIIs) and increased number of antimicrobial prescriptions in patients with PAI compared to population-based matched controls [53]. In patients with AI, prompt treatment of infectious diseases is essential to minimize hospitalization and reduce adrenal crisis [34]. It is also important to emphasize that infections are actually the most common precipitating factor for adrenal crisis, especially gastroenteritis and bronchopulmonary and urinary tract infections [34, 54, 55]. Our results confirmed a greater prevalence and odds of contracting GIIs, MYC and UTI in hypocortisolaemic patients, who also experienced a longer duration of LRTIs.

In this regard, it is still unclear whether the higher risk of infections is correlated with GC replacement therapy doses or regimens in any way. We recently showed that patients with AI exhibit a selective immune impairment, mainly of innate immunity, which can be partially restored by a more circadian rhythm of GC administration [35, 56-58]. This immune impairment was further linked to circadian gene disruption of AI patients [59]. Our study is the first to show that patients with AI have a high risk of infectious disease, regardless of their GC total daily dose. However, in our cohort the median hydrocortisone equivalent dose was 20 mg/day, and patients were not overtreated. Further studies are needed to confirm these findings and to better clarify the contribution of a tailored circadian replacement treatment in AI.

Our study has some limitations. First, caution should be exercised in the interpretation of our results, given that the data were self-reported and recall bias should be taken into account, with some infections possibly being counted by the patient more than once. However, in a shorter version of the ICARO questionnaire used previously in the DREAM trial, we had established that recall bias was minimal [35]. Moreover, the questionnaire was derived from validated questionnaires with a similar time frame and administered in the outpatient clinics of University Hospitals to participants who are used to remembering and answering questions about their conditions and symptoms.

Another limitation is the baseline differences in some demographic characteristics of the hypercortisolism group; this is a common occurrence with observational studies and is a consequence of the nature of this disease. As expected, hypercortisolism was more prevalent in women, and patients more frequently had diabetes and obesity than controls [9]. We also found that the majority of patients with pACS were older, and this is also consistent with previous large studies [49, 50, 60]. To fix all these biases, we adjusted the odds ratio for the main confounders predisposing to infections (age, sex, diabetes, menopause/hypogonadism, obesity) and performed a sensitivity analysis excluding patients with obesity and diabetes.

In view of the scant original data on this topic, we believe this study offers several benefits. It is the first development of an objective tool to quantify the burden of infectious disease in endocrine patients. The aforementioned ISAQ was recently tested in patients with PAI, with unsatisfactory results: this may have been due to the nature of the questionnaire, which was originally designed for severely immunocompromised patients [61]. In contrast, the ICARO questionnaire is derived from both the GNC questionnaire and ISAQ and has been specifically adapted for outpatients. It could thus be a useful tool for future studies aiming to explore the relationships between infection type and severity, hormone levels and treatment of glucocorticoid disorders. The questionnaire proved capable of investigating the epidemiology of the most common infections experienced by patients with endogenous hypercortisolism (excluding ectopic CS), adrenal incidentalomas and pACS, SAI and PAI. Above all, it includes infectious episodes that do not lead to hospitalization or death. These could easily be missed by registry/insurance sources, as patients are less likely to seek medical advice for trivial infections (UTIs, SSTIs or FLU), which however, could still potentially trigger a more dangerous condition. In this context, the self-reported ICARO questionnaire may be a simple, cost-effective new method to: a) identify patients at a higher risk of infections and evaluate the effect of specific treatments on their susceptibility to infection; b) provide important information both for clinicians and patients as to the vulnerability for such infection c) improve patients care and quality of life.

## Data Availability

All data produced in the present work are contained in the manuscript

## Acknowledgements

We thank all patients who participated in the study. We also thank the MCEL group (https://sites.google.com/uniroma1.it/mcel/people) for helping in patient recruitment. Marie-Hélène Hayles MITI revised the English.

## REFERENCES

1. Arlt, W., et al., Endocrinology in the time of COVID-19. Eur J Endocrinol, 2020. 183(1): p. E1–E2.

2. Isidori, A.M., et al., The Italian Endo-COVID-19. Our response to the emergency. J Endocrinol Invest, 2020. 43(7): p. 1039–1041.

3. Arlt, W., et al., ENDOCRINOLOGY IN THE TIME OF COVID-19: Management of adrenal insufficiency. Eur J Endocrinol, 2020. 183(1): p. G25–G32.

4. Isidori, A.M., et al., Use of glucocorticoids in patients with adrenal insufficiency and COVID-19 infection. The Lancet Diabetes & Endocrinology, 2020. 8(6): p. 472–473.

5. Newell-Price, J., et al., ENDOCRINOLOGY IN THE TIME OF COVID-19: Management of Cushing’s syndrome. Eur J Endocrinol, 2020. 183(1): p. G1–G7.

6. Isidori, A.M., et al., COVID-19 infection and glucocorticoids: update from the Italian Society of Endocrinology Expert Opinion on steroid replacement in adrenal insufficiency. J Endocrinol Invest, 2020. 43(8): p. 1141–1147.

7. Guarnotta, V., et al., Glucocorticoid excess and COVID-19 disease. Rev Endocr Metab Disord, 2021. 22(4): p. 703–714.

8. Hasenmajer, V., et al., The Immune System in Cushing’s Syndrome. Trends Endocrinol Metab, 2020. 31(9): p. 655–669.

9. Pivonello, R., et al., Complications of Cushing’s syndrome: state of the art. Lancet Diabetes Endocrinol, 2016. 4(7): p. 611–29.

10. Isidori, A.M., et al., Mechanisms in endocrinology: The spectrum of haemostatic abnormalities in glucocorticoid excess and defect. Eur J Endocrinol, 2015. 173(3): p. R101–13.

11. Isidori, A.M., et al., The hypertension of Cushing’s syndrome: controversies in the pathophysiology and focus on cardiovascular complications. J Hypertens, 2015. 33(1): p. 44–60.

12. Clayton, R.N., Mortality in Cushing’s disease. Neuroendocrinology, 2010. 92 Suppl 1: p. 71–6.

13. Dekkers, O.M., et al., Multisystem morbidity and mortality in Cushing’s syndrome: a cohort study. J Clin Endocrinol Metab, 2013. 98(6): p. 2277–84.

14. Ragnarsson, O., et al., Overall and Disease-Specific Mortality in Patients With Cushing Disease: A Swedish Nationwide Study. J Clin Endocrinol Metab, 2019. 104(6): p. 2375–2384.

15. Valassi, E., et al., High mortality within 90 days of diagnosis in patients with Cushing’s syndrome: results from the ERCUSYN registry. Eur J Endocrinol, 2019. 181(5): p. 461–472.

16. Sarlis, N.J., S.J. Chanock, and L.K. Nieman, Cortisolemic indices predict severe infections in Cushing syndrome due to ectopic production of adrenocorticotropin. J Clin Endocrinol Metab, 2000. 85(1): p. 42–7.

17. Ilias, I., et al., Cushing’s syndrome due to ectopic corticotropin secretion: twenty years’ experience at the National Institutes of Health. J Clin Endocrinol Metab, 2005. 90(8): p. 4955–62.

18. Sathyakumar, S., et al., Ectopic Cushing Syndrome: A 10-Year Experience from a Tertiary Care Center in Southern India. Endocr Pract, 2017. 23(8): p. 907–914.

19. Ejaz, S., et al., Cushing syndrome secondary to ectopic adrenocorticotropic hormone secretion: the University of Texas MD Anderson Cancer Center Experience. Cancer, 2011. 117(19): p. 4381–9.

20. Fassnacht, M., et al., Management of adrenal incidentalomas: European Society of Endocrinology Clinical Practice Guideline in collaboration with the European Network for the Study of Adrenal Tumors. Eur J Endocrinol, 2016. 175(2): p. G1–G34.

21. Sbardella, E., et al., Cardiovascular features of possible autonomous cortisol secretion in patients with adrenal incidentalomas. Eur J Endocrinol, 2018. 178(5): p. 501–511.

22. Debono, M. and J. Newell-Price, Subclinical hypercortisolism in adrenal incidentaloma. Curr Opin Endocrinol Diabetes Obes, 2015. 22(3): p. 185–92.

23. Chiodini, I., et al., MECHANISMS IN ENDOCRINOLOGY: Endogenous subclinical hypercortisolism and bone: a clinical review. Eur J Endocrinol, 2016. 175(6): p. R265–R282.

24. Bergthorsdottir, R., et al., Premature mortality in patients with Addison’s disease: a population-based study. J Clin Endocrinol Metab, 2006. 91(12): p. 4849–53.

25. Chantzichristos, D., et al., Mortality in patients with diabetes mellitus and Addison’s disease: a nationwide, matched, observational cohort study. Eur J Endocrinol, 2017. 176(1): p. 31–39.

26. Erichsen, M.M., et al., Normal overall mortality rate in Addison’s disease, but young patients are at risk of premature death. Eur J Endocrinol, 2009. 160(2): p. 233–7.

27. Quinkler, M., et al., Mortality data from the European Adrenal Insufficiency Registry-Patient characterization and associations. Clin Endocrinol (Oxf), 2018. 89(1): p. 30–35.

28. Mills, J.L., et al., Long-term mortality in the United States cohort of pituitary-derived growth hormone recipients. J Pediatr, 2004. 144(4): p. 430–6.

29. Burman, P., et al., Deaths among adult patients with hypopituitarism: hypocortisolism during acute stress, and de novo malignant brain tumors contribute to an increased mortality. J Clin Endocrinol Metab, 2013. 98(4): p. 1466–75.

30. Smans, L.C., et al., Increased use of antimicrobial agents and hospital admission for infections in patients with primary adrenal insufficiency: a cohort study. Eur J Endocrinol, 2013. 168(4): p. 609–14.

31. Bjornsdottir, S., et al., Drug prescription patterns in patients with Addison’s disease: a Swedish population-based cohort study. J Clin Endocrinol Metab, 2013. 98(5): p. 2009–18.

32. Hahner, S., et al., High incidence of adrenal crisis in educated patients with chronic adrenal insufficiency: a prospective study. J Clin Endocrinol Metab, 2015. 100(2): p. 407–16.

33. Martin-Grace, J., et al., Adrenal insufficiency: Physiology, clinical presentation and diagnostic challenges. Clin Chim Acta, 2020. 505: p. 78–91.

34. Smans, L.C., et al., Incidence of adrenal crisis in patients with adrenal insufficiency. Clin Endocrinol (Oxf), 2016. 84(1): p. 17–22.

35. Isidori, A.M., et al., Effect of once-daily, modified-release hydrocortisone versus standard glucocorticoid therapy on metabolism and innate immunity in patients with adrenal insufficiency (DREAM): a single-blind, randomised controlled trial. Lancet Diabetes Endocrinol, 2018. 6(3): p. 173–185.

36. Sievers, C., et al., Evaluation of a questionnaire to assess selected infectious diseases and their risk factors : findings of a multicenter study. Bundesgesundheitsblatt Gesundheitsforschung Gesundheitsschutz, 2014. 57(11): p. 1283–91.

37. Castell, S., et al., Test-retest reliability of an infectious disease questionnaire and evaluation of self-assessed vulnerability to infections : findings of Pretest 2 of the German National Cohort. Bundesgesundheitsblatt Gesundheitsforschung Gesundheitsschutz, 2014. 57(11): p. 1300–7.

38. Peter, H.-H., Construction and Clinical Validation of a Questionnaire-based Risk Score to Identify Patients Suffering from Immunodeficiency or Systemic Autoimmunity. British Journal of Medicine and Medical Research, 2014. 4(29): p. 4751–4769.

39. Nieman, L.K., et al., The diagnosis of Cushing’s syndrome: an Endocrine Society Clinical Practice Guideline. J Clin Endocrinol Metab, 2008. 93(5): p. 1526–40.

40. Fleseriu, M., et al., Consensus on diagnosis and management of Cushing’s disease: a guideline update. Lancet Diabetes Endocrinol, 2021. 9(12): p. 847–875.

41. Bornstein, S.R., et al., Diagnosis and Treatment of Primary Adrenal Insufficiency: An Endocrine Society Clinical Practice Guideline. J Clin Endocrinol Metab, 2016. 101(2): p. 364–89.

42. Osborne Jason W.C.A.B., Sample size and subject to item ratio in principal components analysis.. Practical Assessment, Research, and Evaluation 2004. 9: p. 8.

43. Groenwold, R.H.H., et al., Multiple testing: when is many too much? Eur J Endocrinol, 2021. 184(3): p. E11–E14.

44. Plotz, C.M., A.I. Knowlton, and C. Ragan, The natural history of Cushing’s syndrome. Am J Med, 1952. 13(5): p. 597–614.

45. Broder, M.S., et al., Incremental healthcare resource utilization and costs in US patients with Cushing’s disease compared with diabetes mellitus and population controls. Pituitary, 2015. 18(6): p. 796–802.

46. Scaroni, C., et al., Approach to patients with pseudo-Cushing’s states. Endocr Connect, 2020. 9(1): p. R1–R13.

47. Bancos, I. and A. Prete, Approach to the Patient With Adrenal Incidentaloma. J Clin Endocrinol Metab, 2021. 106(11): p. 3331–3353.

48. Di Dalmazi, G., Adrenal Incidentaloma: Picking out the High-Risk Patients. Exp Clin Endocrinol Diabetes, 2019. 127(2-03): p. 178–184.

49. Morelli, V., et al., Long-term follow-up in adrenal incidentalomas: an Italian multicenter study. J Clin Endocrinol Metab, 2014. 99(3): p. 827–34.

50. Debono, M., et al., Cortisol as a marker for increased mortality in patients with incidental adrenocortical adenomas. J Clin Endocrinol Metab, 2014. 99(12): p. 4462–70.

51. Sharma, M.D., et al., High frequency of serious infections in patients with panhypopituitarism: a case-control study. Clin Infect Dis, 2001. 32(1): p. 153–8.

52. Tomlinson, J.W., et al., Association between premature mortality and hypopituitarism. West Midlands Prospective Hypopituitary Study Group. Lancet, 2001. 357(9254): p. 425–31.

53. Tresoldi, A.S., et al., Increased Infection Risk in Addison’s Disease and Congenital Adrenal Hyperplasia. J Clin Endocrinol Metab, 2020. 105(2).

54. Quinkler, M., et al., Characterization of patients with adrenal insufficiency and frequent adrenal crises. Eur J Endocrinol, 2021. 184(6): p. 761–771.

55. Hahner, S., et al., Epidemiology of adrenal crisis in chronic adrenal insufficiency: the need for new prevention strategies. Eur J Endocrinol, 2010. 162(3): p. 597–602.

56. Isidori, A.M., et al., Towards the tailoring of glucocorticoid replacement in adrenal insufficiency: the Italian Society of Endocrinology Expert Opinion. J Endocrinol Invest, 2020. 43(5): p. 683–696.

57. Minnetti, M., et al., Fixing the broken clock in adrenal disorders: focus on glucocorticoids and chronotherapy. J Endocrinol, 2020. 246(2): p. R13–R31.

58. Bancos, I., et al., Primary adrenal insufficiency is associated with impaired natural killer cell function: a potential link to increased mortality. Eur J Endocrinol, 2017. 176(4): p. 471–480.

59. Venneri, M.A., et al., Circadian Rhythm of Glucocorticoid Administration Entrains Clock Genes in Immune Cells: A DREAM Trial Ancillary Study. J Clin Endocrinol Metab, 2018. 103(8): p. 2998–3009.

60. Schalin-Jantti, C., et al., A 5-Year Prospective Follow-Up Study of Lipid-Rich Adrenal Incidentalomas: No Tumor Growth or Development of Hormonal Hypersecretion. Endocrinol Metab (Seoul), 2015. 30(4): p. 481–7.

61. Quinkler, M. and T. Kienitz, The ISAQ Score Does Not Predict Adrenal Crisis in Patients with Primary Adrenal Insufficiency. Exp Clin Endocrinol Diabetes, 2022.

