## Supplementary Data for "Susceptibility and characteristics of infections in patients with glucocorticoid excess or insufficiency: The ICARO tool"

**Supplementary material:** self-administered questionnaire completed by the study participants

**ICARO questionnaire (InfeCtions in pAtients with endocRinOpathies).** The following questions assess the frequency, duration and treatment of any infections you have had in the past 12 months. Please add up how often you suffered from these infections. For example if you had a cold 2 times and a middle ear infection once, then you had 3 upper respiratory tract infections (so you should choose the option "3 or 4 times"). A clinician is available for any questions or clarifications on the questionnaire.

**How often have you had an infection of the upper respiratory tract (e.g. cold or an infection of the sinus, tonsils, middle ear (otitis media), throat, larynx, pharynx) <u>in the past 12 months?</u>**

| None (0.5) | 1 or 2 times (2) | 3 or 4 times (4) | 5 or 6 times (4) | more than 6 times (4) |
|---|---|---|---|---|
| ☐ | ☐ | ☐ | ☐ | ☐ |

| **How long did each infectious episode last?** | | | | **Did you take any medicines to treat the infection?** | | |
|---|---|---|---|---|---|---|
| less than one week (0) | from one to two weeks (2) | from two to three weeks (4) | more than three weeks (4) | no | yes, antibiotics | yes, antifungals |
| ☐ | ☐ | ☐ | ☐ | ☐ | ☐ | ☐ |

**How often have you had bronchitis or pneumonia <u>in the past 12 months</u>?**

| None (0.5) | 1 or 2 times (2) | 3 or 4 times (4) | 5 or 6 times (4) | more than 6 times (4) |
|---|---|---|---|---|
| ☐ | ☐ | ☐ | ☐ | ☐ |

| **How long did each infectious episode last?** | | | | **Did you take any medicines to treat the infection?** | | |
|---|---|---|---|---|---|---|
| less than one week (0) | from one to two weeks (2) | from two to three weeks (4) | more than three weeks (4) | no | yes, antibiotics | yes, antivirals |
| ☐ | ☐ | ☐ | ☐ | ☐ | ☐ | ☐ |

**How often have you had a gastrointestinal infection (stomach bug) <u>in the past 12 months?</u>**

| None (0.5) | 1 or 2 times (2) | 3 or 4 times (4) | 5 or 6 times (4) | more than 6 times (4) |
|---|---|---|---|---|
| ☐ | ☐ | ☐ | ☐ | ☐ |

| **How long did each infectious episode last?** | | | | **Did you take any medicines to treat the infection?** | |
|---|---|---|---|---|---|
| less than one week (0) | from one to two weeks (2) | from two to three weeks (4) | more than three weeks (4) | no | yes, antibiotics |
| ☐ | ☐ | ☐ | ☐ | ☐ | ☐ |

**How often have you had a skin or soft tissue infection <u>in the past 12 months</u>? (Herpes labialis (cold sore), warts, herpes zoster (chickenpox/shingles), conjunctivitis)**

| None (0.5) | 1 or 2 times (2) | 3 or 4 times (4) | 5 or 6 times (4) | more than 6 times (4) |
|---|---|---|---|---|
| ☐ | ☐ | ☐ | ☐ | ☐ |

| **How long did each infectious episode last?** | | | | **Did you take any medicines to treat the infection?** | | |
|---|---|---|---|---|---|---|
| less than one week | from one to two weeks | from two to three weeks | more than three weeks | no | yes, antibiotics | yes, antivirals |
| ☐ (0) | ☐ (2) | ☐ (4) | ☐ (4) | ☐ | ☐ | ☐ |

**How often have you had a skin or soft tissue infection <u>in the past 12 months</u>? (boils, abscesses, styes)**

| None (0.5) | 1 or 2 times (2) | 3 or 4 times (4) | 5 or 6 times (4) | more than 6 times (4) |
|---|---|---|---|---|
| ☐ | ☐ | ☐ | ☐ | ☐ |

| **How long did each infectious episode last?** | | | | **Did you take any medicines to treat the infection?** | |
|---|---|---|---|---|---|
| less than one week (0) | from one to two weeks (2) | from two to three weeks (4) | more than three weeks (4) | no | yes, antibiotics |
| ☐ | ☐ | ☐ | ☐ | ☐ | ☐ |

**How often have you had a fungal infection (e.g. fungal infection of the fingernails or toenails, oral thrush, vaginal thrush) <u>in the past 12 months</u>?**

| None (0.5) | 1 or 2 times (2) | 3 or 4 times (4) | 5 or 6 times (4) | more than 6 times (4) |
|---|---|---|---|---|
| ☐ | ☐ | ☐ | ☐ | ☐ |

| **How long did each infectious episode last?** | | | | **Did you take any medicines to treat the infection?** | |
|---|---|---|---|---|---|
| less than one | from one to two | from two to | more than three | no | yes, antifungals |

| | | | | | |
|---|---|---|---|---|---|
| week (0) | weeks (2) | three weeks (4) | weeks (4) | | |
| ☐ | ☐ | ☐ | ☐ | ☐ | ☐ |

| | | | | |
|---|---|---|---|---|
| **How often have you had a genital infection (e.g. chlamydia, gonorrhea, syphilis) <u>in the past 12 months</u>?** | | | | |
| None (0.5) | 1 or 2 times (2) | 3 or 4 times (4) | 5 or 6 times (4) | more than 6 times (4) |
| ☐ | ☐ | ☐ | ☐ | ☐ |

| | | | | | |
|---|---|---|---|---|---|
| **How long did each infectious episode last?** | | | | **Did you take any medicines to treat the infection?** | |
| less than one week (0) | from one to two weeks (2) | from two to three weeks (4) | more than three weeks (4) | no | yes, antibiotics |
| ☐ | ☐ | ☐ | ☐ | ☐ | ☐ |

| | | | | |
|---|---|---|---|---|
| **How often have you had a lower urinary tract infection (e.g. bladder infection) <u>in the past 12 months</u>?** | | | | |
| None (0.5) | 1 or 2 times (2) | 3 or 4 times (4) | 5 or 6 times (4) | more than 6 times (4) |
| ☐ | ☐ | ☐ | ☐ | ☐ |

| | | | | | | |
|---|---|---|---|---|---|---|
| **How long did each infectious episode last?** | | | | **Did you take any medicines to treat the infection?** | | |
| less than one week (0) | from one to two weeks (2) | from two to three weeks (4) | more than three weeks (4) | no | yes, antibiotics | yes, antifungals |
| ☐ | ☐ | ☐ | ☐ | ☐ | ☐ | ☐ |

| | | | | |
|---|---|---|---|---|
| **How often have you had the flu <u>in the past 12 months</u>?** | | | | |
| None (0.5) | 1 or 2 times (2) | 3 or 4 times (4) | 5 or 6 times (4) | more than 6 times (4) |
| ☐ | ☐ | ☐ | ☐ | ☐ |

| | |
|---|---|
| **Have you been hospitalized for infectious diseases <u>in the past 12 months</u>?** | |
| no | yes |
| ☐ | ☐ |

| | | | | |
|---|---|---|---|---|
| **Have you been vaccinated in the past year?** | | **If so, which vaccines did you get in the past year?** | | |
| yes | no | flu | pneumococcus | other* |
| ☐ | ☐ | ☐ | ☐ | ☐ |
| | | *, specify:........................................................................................ | | |

| | | | |
|---|---|---|---|
| **Have you been absent from work due to infectious disease?** | | **If yes, how long?** | |
| Yes | No | Less than 7 days | More than 7 days |
| ☐ | ☐ | ☐ | ☐ |

**Supplementary table 1.** Duration of infectious diseases in patients with hypocortisolism having the infection in the last 12 months. Categorical variables are expressed as percentages and frequencies. p* values for comparisons with controls. $p^{\S}$ values for comparisons between CS and pACS. After correction for multiple testing a $p < 0.05$ was considered as significant statistically different (bold). [a,b,c,d,e,f,g,h,i]: significantly different duration respect to controls at post hoc analysis using multiple z-tests with Bonferroni correction.

| | Controls (n=570) | Hypercortisolism (n=75) | *p** | CS (N=43) | *p** | pACS (N=32) | *p** | $p^{\S}$ |
|---|---|---|---|---|---|---|---|---|
| **Upper Respiratory tract infections (URTis) duration** | | | | | | | | |
| less than 1 week | 65%(279/429) | 60%(38/63) | | 66%(23/35) | | 54%(15/28) | | |
| from 1 to 2 weeks | 28%(122/429) | 32%(20/63) | 0.743 | 26%(9/35) | 0.903 | 39%(11/28) | 0.742 | 0.516 |
| from 2 to 3 weeks | 4%(19/429) | 8%(5/63) | | 9%(3/35) | | 7%(2/28) | | |
| more than 3 weeks | 2%(9/429) | 0%(0) | | 0%(0) | | 0%(0) | | |
| **Lower respiratory tract infections (LRTis) duration** | | | | | | | | |
| less than 1 week | 30%(19/64) | 11%(2/18) | | 10%(1/10) | | 12.5%(1/8) | | |
| from 1 to 2 weeks | 55%(35/64) | 72%(13/18) | 0.747 | 60%(6/10) | 0.060 | 87.5%(7/8) | 0.742 | 0.409 |
| from 2 to 3 weeks | 6%(3/64) | 11%(2/18) | | 20%(2/10) | | 0%(0) | | |
| more than 3 weeks | 9%(6/64) | 6%(1/18) | | 10%(1/10) | | 0%(0) | | |
| **Gastrointestinal tract infections (GIIs) duration** | | | | | | | | |
| less than 1 week | 88%(163/185)[a] | 58%(18/31)[a] | | 57%(12/21)[a] | | 65%(11/17) | | |
| from 1 to 2 weeks | 11%(20/185)[b] | 29%(9/31)[b] | **0.029** | 29%(6/21)[b] | **0.037** | 29%(5/17) | 0.114 | 0.946 |
| from 2 to 3 weeks | 0.5%(1/185) | 0%(0) | | 0%(0) | | 0%(0) | | |
| more than 3 weeks | 0.5%(1/185)[c] | 13%(4/31)[c] | | 14%(3/21)[c] | | 6%(1/17) | | |
| **Skin infections 1 (SSTis-1) duration** | | | | | | | | |
| less than 1 week | 56%(79/142) | 61%(11/18) | | 50%(5/10) | | 75%(6/8) | | |
| from 1 to 2 weeks | 30%(42/142) | 22%(4/18) | 1.000 | 20%(2/10) | 0.790 | 25%(2/8) | 0.901 | 0.407 |
| from 2 to 3 weeks | 6%(9/142) | 11%(2/18) | | 20%(2/10) | | 0%(0) | | |
| more than 3 weeks | 8%(12/142) | 6%(1/18) | | 10%(1/10) | | 0%(0) | | |
| **Skin infections 2 (SSTis-2) duration** | | | | | | | | |
| less than 1 week | 56%(49/88)[d] | 29%(4/14) | | 20%(2/10)[d] | | 50%(2/4) | | |
| from 1 to 2 weeks | 33%(29/88) | 29%(4/14) | 0.342 | 30%(3/10) | **0.034** | 25%(1/4) | 0.290 | 0.540 |
| from 2 to 3 weeks | 2%(2/88)[e] | 21%(3/14) | | 20%(2/10)[e] | | 25%(1/4) | | |
| more than 3 weeks | 9%(8/88)[f] | 21%(3/14) | | 30%(3/10)[f] | | 0%(0) | | |
| **Mycosis (MYC) duration** | | | | | | | | |
| less than 1 week | 33%(21/63)[g] | 39%(9/23)[g] | | 33%(6/18)[g] | | 60%(3/5) | | |
| from 1 to 2 weeks | 48%(30/63)[h] | 13%(3/23)[h] | **0.025** | 6%(1/18)[h] | **0.016** | 40%(2/5) | 0.894 | 0.062 |
| from 2 to 3 weeks | 5%(3/63)[i] | 26%(6/23)[i] | | 33%(6/18)[i] | | 0%(0) | | |
| more than 3 weeks | 14%(9/63) | 22%(5/23) | | 28%(5/18) | | 0%(0) | | |
| **Sexually transmitted infections (STIs) duration** | | | | | | | | |
| less than 1 week | 22%(2/9) | 100%(2/2) | | 100%(1/1) | | 100%(1/1) | | |
| from 1 to 2 weeks | 67%(6/9) | 0%(0) | 0.359 | 0%(0) | 0.651 | 0%(0) | 0.644 | 0.143 |
| from 2 to 3 weeks | 0% (0) | 0%(0) | | 0%(0) | | 0%(0) | | |
| more than 3 weeks | 11%(1/9) | 0%(0) | | 0%(0) | | 0%(0) | | |
| **Urinary tract infections (UTIs) duration** | | | | | | | | |
| less than 1 week | 70%(59/86) | 59.5%(22/37) | | 56.5%(13/23) | | 64%(9/14) | | |
| from 1 to 2 weeks | 24%(21/86) | 22%(8/37) | 0.641 | 26%(6/23) | 0.802 | 14%(2/14) | 0.758 | 0.843 |
| from 2 to 3 weeks | 2%(2/86) | 5%(2/37) | | 4%(1/23) | | 7%(1/14) | | |
| More than 3 weeks | 5%(4/86) | 13.5%(5/37) | | 13%(3/23) | | 14%(2/14) | | |

**Supplementary Table 2.** duration of infectious diseases in patients with hypocortisolism. Categorical variables are expressed as percentages and frequencies. *p** values for comparisons with controls. $p^{§}$ values for comparisons between PAI and SAI. After correction for multiple testing a $p < 0.05$ was considered as significant statistically different (bold). [a,b.c] significantly different duration respect to controls at post hoc analysis using multiple z-tests with Bonferroni correction.

| | **Controls** (n=570) | **Hypocortisolism** (n=135) | *p** | **PAI** (n=32) | *p** | **SAI** (n=103) | *p** | $P^{§}$ |
|---|---|---|---|---|---|---|---|---|
| **Upper Respiratory tract infections (URTis) duration** | | | | | | | | |
| less than 1 week | 65%(279/429)[a] | 53%(55/103) | 0.459 | 35%(8/23)[a] | **0.036** | 59%(47/80) | 0.748 | 0.058 |
| from 1 to 2 weeks | 28%(122) | 39%(40) | | 45.5%(10) | | 37.5%(30) | | |
| from 2 to 3 weeks | 4%(19) | 5%(5) | | 13%(3) | | 2.5%(2) | | |
| more than 3 weeks | 2%(9)[b] | 3%(3) | | 8.5%(2)[b] | | 1%(1) | | |
| **Lower respiratory tract infections (LRTis) duration** | | | | | | | | |
| less than 1 week | 30%(19/64) | 7%(1) | **0.033** | 33%(2/6) | 0.291 | 9%(1/11) | 0.143 | 0.611 |
| from 1 to 2 weeks | 55%(35) | 33%(5) | | 17%(1) | | 36%(4) | | |
| from 2 to 3 weeks | 6%(3)[c] | 35%(6)[c] | | 33%(2) | | 36%(4) | | |
| more than 3 weeks | 9%(6) | 18%(3) | | 17%(1) | | 18%(2) | | |
| **Gastrointestinal tract infections (GIIs) duration** | | | | | | | | |
| less than 1 week | 88%(163/185) | 75%(51/68) | 0.200 | 65%(11/17) | 0.083 | 65%(11/17) | 0.736 | 0.161 |
| from 1 to 2 weeks | 11%(20) | 23.5%(13) | | 29%(5) | | 29%(5) | | |
| from 2 to 3 weeks | 0.5%(1) | 0%(0) | | 0%(0) | | 0%(0) | | |
| more than 3 weeks | 0.5%(1) | 1.5%(1) | | 6%(1) | | 6%(1) | | |
| **Skin infections 1 (SSTis-1) duration** | | | | | | | | |
| less than 1 week | 56%(79/142) | 50%(19/38) | 1.000 | 38.5%(5/13) | 0.469 | 9%(1/11) | 0.141 | 0.098 |
| from 1 to 2 weeks | 30%(42) | 32%(12) | | 23%(3) | | 36%(4) | | |
| from 2 to 3 weeks | 6%(9) | 5%(2) | | 15%(2) | | 36%(4) | | |
| more than 3 weeks | 8%(12) | 13%(5) | | 23%(3) | | 18%(0) | | |
| **Skin infections 2 (SSTis-2) duration** | | | | | | | | |
| less than 1 week | 56%(49/88) | 67%(12/18) | 0.795 | 100%(3/3) | 0.844 | 56%(14/25) | 0.879 | 0.407 |
| from 1 to 2 weeks | 33%(29) | 28%(5) | | 0%(0) | | 36%(9) | | |
| from 2 to 3 weeks | 2%(2) | 5%(1) | | 0%(0) | | 0%(1) | | |
| more than 3 weeks | 9%(8) | 0%(0) | | 0%(0) | | 8%(2) | | |
| **Mycosis (MYC) duration** | | | | | | | | |
| less than 1 week | 33%(21/63) | 33%(10/30) | 0.076 | 43%(3/7) | 0.893 | 30%(7/23) | 0.228 | 0.052 |
| from 1 to 2 weeks | 48%(30) | 10%(3) | | 29%(2) | | 4%(1) | | |
| from 2 to 3 weeks | 5%(3) | 40%(12) | | 0%(0) | | 52%(12) | | |
| more than 3 weeks | 14%(9) | 17%(5) | | 29%(2) | | 13%(3) | | |
| **Sexually transmitted infections (STIs) duration** | | | | | | | | |
| less than 1 week | 22%(2/9) | 33%(1/3) | 0.547 | 0%(0/1) | 0.347 | 50%(1/2) | 0.897 | 0.223 |
| from 1 to 2 weeks | 67%(6) | 33%(1) | | 0%(0) | | 50%(1) | | |
| from 2 to 3 weeks | 0% (0) | 33%(1) | | 0%(0) | | 0%(0) | | |
| more than 3 weeks | 11%(1) | 33%(1) | | 100%(1) | | 0%(0) | | |
| **Urinary tract infections (UTIs) duration** | | | | | | | | |
| less than 1 week | 70%(59/86) | 73%(30/41) | 1.000 | 75%(9/12) | 1.000 | 72%(21/29) | 1.000 | 0.824 |
| from 1 to 2 weeks | 24%(21) | 22%(9) | | 25%(3) | | 21%(6) | | |
| from 2 to 3 weeks | 2%(2) | 2.5%(1) | | 20%(0) | | 3%(1) | | |
| more than 3 weeks | 5%(4) | 2.5%(1) | | 0%(0) | | 3%(1) | | |
